# Detecting short-interval longitudinal cortical atrophy in neurodegenerative dementias via cluster scanning: A proof of concept

**DOI:** 10.1101/2025.03.14.25323769

**Authors:** Yuta Katsumi, Michael Brickhouse, Lindsay C. Hanford, Jared A. Nielsen, Maxwell L. Elliott, Ross W. Mair, Alexandra Touroutoglou, Mark C. Eldaief, Randy L. Buckner, Bradford C. Dickerson

**Author notes:** Corresponding authors: Yuta Katsumi, PhD and Brad Dickerson, MD. Equal contributions (Yuta Katsumi, Michael Brickhouse, Randy L. Buckner, Bradford C. Dickerson).

## Abstract

Regional brain atrophy estimated from structural magnetic resonance imaging (MRI) is a widely used measure of neurodegeneration in Alzheimer’s disease (AD), Frontotemporal Lobar Degeneration (FTLD), and other dementias. Yet, traditional MRI-derived morphometric estimates are susceptible to measurement errors, posing a challenge for reliably detecting longitudinal atrophy, particularly over short intervals. Here, we examined the utility of multiple MRI scans acquired in rapid succession (i.e., *cluster scanning*) for detecting longitudinal cortical atrophy over 3- and 6-month intervals within individual patients. Four individuals with mild cognitive impairment or mild dementia likely due to AD or FTLD participated in this study. At baseline, 3 months, and 6 months, structural MRI data were collected on a 3 Tesla scanner using a fast 1.2-mm T1-weighted multi-echo magnetization-prepared rapid gradient echo (MEMPRAGE) sequence (acquisition time = 2’23’’). At each timepoint, participants underwent up to 32 MEMPRAGE scans acquired in four separate sessions over two days. Using linear mixed-effects models, phenotypically vulnerable cortical (“core atrophy”) regions exhibited statistically significant longitudinal atrophy in all participants (i.e., decreased cortical thickness) by 3 months and further demonstrated preferential vulnerability compared to control regions in three of the participants over at least one of the 3-month intervals. These findings provide proof-of-concept evidence that pooling multiple morphometric estimates derived from cluster scanning can detect longitudinal cortical atrophy over short intervals in individual patients with neurodegenerative dementias.

## 1. Introduction

Regional brain atrophy estimated from structural magnetic resonance imaging (MRI) is a widely used measure of neurodegeneration in Alzheimer’s disease (AD), Frontotemporal Lobar Degeneration (FTLD), and other dementias. There is now ample evidence to suggest that MRI-derived brain atrophy estimates are useful for diagnosis, prognostication, and longitudinal outcome monitoring in neurodegenerative disease. For example, we and others previously identified spatially distinct “signature” patterns of cortical atrophy in early symptomatic patients with late-onset AD (Dickerson et al., 2009; Jack et al., 2015; Jack & Holtzman, 2013), early-onset AD (Frisoni et al., 2007; Harper et al., 2017; Möller et al., 2013; Touroutoglou et al., 2023) and various clinical phenotypes of FTLD (Agosta et al., 2015; Caso et al., 2014; Eldaief et al., 2023; Harper et al., 2017; Rohrer et al., 2011; Seeley et al., 2008; Vemuri et al., 2011; Zhang et al., 2013), which are highly replicable across independent samples and robustly associated with symptom severity. Regional atrophy patterns are less robust and consistent in Dementia with Lewy bodies (Harper et al., 2017; Vemuri et al., 2011; Ye et al., 2020). The magnitude of MRI-based cortical atrophy in these signature regions and other phenotypically vulnerable brain networks can predict subsequent clinical progression or decline among individuals at early clinical stages of AD or FTLD (Anderl-Straub et al., 2021; Bakkour et al., 2009; Dickerson & Wolk, 2013; Katsumi et al., 2023, 2024; Keret et al., 2021; Staffaroni et al., 2019) as well as individuals who develop these syndromes but were scanned when cognitively unimpaired (Dickerson et al., 2011; Prosser et al., 2023). Longitudinal MRI studies have demonstrated unique spatiotemporal characteristics of brain atrophy progression across phenotypic subtypes of AD and FTLD, while also identifying the relationship between the pattern of longitudinal atrophy and other imaging biomarkers of neuropathologic change (Bejanin et al., 2020; Harrison et al., 2019; La Joie et al., 2020; Phillips et al., 2019; Ranasinghe et al., 2021; Sintini et al., 2019, 2020). In addition to its utility for morphometric analyses, structural MRI is commonly used as an anatomical reference for other neuroimaging modalities such as positron emission tomography (PET), functional MRI, or diffusion MRI data, further highlighting the importance of obtaining reliable morphometrics for the accurate characterization of each patient’s brain structural integrity.

Traditional MRI measures of brain atrophy can reveal submillimeter structural abnormalities in neurodegenerative patients. However, MRI-derived morphometric estimates are prone to small-magnitude measurement errors, which pose a significant challenge for reliably detecting longitudinal brain structural change within an individual patient, particularly over short time intervals. Common morphometric estimates (e.g., regional cortical thickness generated by FreeSurfer) in healthy participants have measurement errors of 2-5%, calculated based on repeat scans acquired during the same imaging session (Tustison et al., 2014) or in sessions a few weeks apart (Dickerson et al., 2008; Han et al., 2006; Jovicich et al., 2009; Wonderlick et al., 2009). The annual rate of atrophy in the hippocampus and late-onset AD signature cortical regions is ∼2-6% in symptomatic AD patients (Barnes et al., 2009; Fjell et al., 2009; Schuff et al., 2009). Therefore, the magnitude of longitudinal atrophy over one year is comparable on average to that of measurement errors. Measuring brain atrophy over short time intervals (< 1 year) has important implications for clinical trial design, as it would allow neurodegeneration to be monitored in smaller, early-phase therapeutic trials, particularly those targeting patients with relatively rapidly progressive forms of dementia (e.g., early-onset AD, behavioral variant FTD) (Lima-Silva et al., 2021; Mendez, 2019). However, the feasibility of reliably estimating short-term atrophy is limited using conventional MRI scans, as most contemporary longitudinal studies of neurodegenerative dementias collect only one or two of these scans annually, such as the Alzheimer’s Disease Neuroimaging Initiative (ADNI) (Veitch et al., 2023), the Dominantly Inherited Alzheimer Network (DIAN) (Moulder et al., 2013), the ARTFL LEFFTDS Longitudinal Frontotemporal Lobar Degeneration study (ALLFTD) (Boeve et al., 2020), the Longitudinal Early-onset Alzheimer’s Disease Study (LEADS) (Apostolova et al., 2021), and the Parkinson Progression Marker Initiative (PPMI) (Marek et al., 2011). Point estimates based on single MRI acquisitions are subject to normal variation around the central tendency, which may lead to over- or underestimation of atrophy rates over time in any given individual (**Figure 1**).

**Figure 1.**
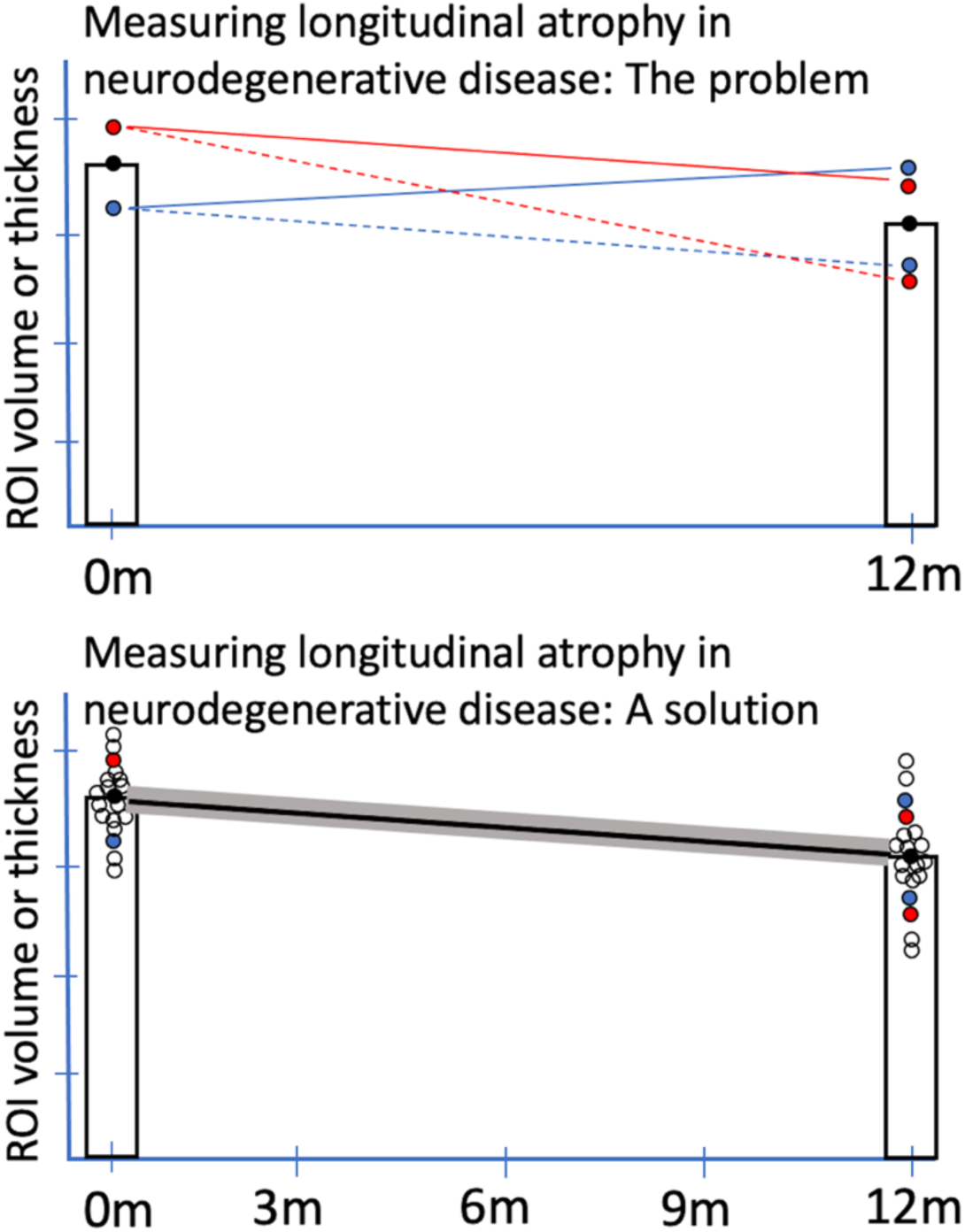
Detection of longitudinal atrophy in neurodegenerative disease. Estimates of longitudinal atrophy rates in neurodegenerative disease are imprecise with current methods, primarily because of undersampling. The top graph illustrates the problem. At each timepoint, a single T1-weighted (T1w) structural MRI scan is typically acquired. It is clear from a variety of studies, including our own, that various sources of error contribute to a distribution of values around the central tendency (represented by black dot). If the baseline estimate is high (red dot), the annualized atrophy rate may be estimated reasonably well at 12-month follow-up (solid red line) or overestimated (dashed red line). If the baseline estimate is low (blue dot), the atrophy rate may be estimated reasonably well (dashed blue line) or may be grossly underestimated (solid blue line). The bottom graph illustrates a proposed solution based on cluster scanning. Dense sampling via cluster scanning at each timepoint should enable not only more precise estimates at typical intervals for longitudinal scanning, but also potentially more sensitive detection of atrophy at shorter intervals, including 6-month or even 3-month intervals.

To improve brain morphometric precision, we have previously introduced the concept of *cluster scanning*, which involves pooling estimates by taking the average of multiple repeated measurements acquired from an individual at essentially one timepoint (Elliott et al., 2024; Nielsen et al., 2019). While still uncommon in structural neuroimaging, achieving greater measurement precision through repeated sampling and data aggregation is an established approach in functional neuroimaging (Birn et al., 2013; Elliott et al., 2019; Gordon et al., 2017; Laumann et al., 2015), psychophysiology (Boudewyn et al., 2018; Huffmeijer et al., 2014), and psychometrics (Crocker & Algina, 1986; Kuder & Richardson, 1937; Nicosia et al., 2023). The current standard for brain structural MRI is a T1w magnetization-prepared rapid gradient echo (MPRAGE) sequence with 1.0 mm isotropic resolution that takes ∼5-7 min to collect on a 3 Tesla scanner. Acquisition of many of these lengthy standard MPRAGE scans per session would be costly and burdensome for participants, especially among symptomatic patients. To achieve dense morphometric sampling within an individual, we have capitalized on extremely rapid MRI sequences with 1.0 mm isotropic resolution, including a compressed-sensing acquisition with six-fold acceleration (CS; acquisition time = 1’12’’) (Mussard et al., 2020) and an acquisition with wave-controlled aliasing in parallel imaging and 3 × 3 acceleration (WAVEx9; acquisition time = 1’9’’) (Polak et al., 2018). Despite the five-fold reduction in acquisition time, we found that a single scan collected with these sequences produces morphometric estimates with high test-retest reliability, high convergent validity, and measurement error rate comparable to those obtained from a standard MPRAGE scan from the ADNI (acquisition time = 5’12’’) (Elliott et al., 2023). In a recent study employing the cluster scanning protocol, we compared brain morphometrics estimated from a single standard MPRAGE scan using the ADNI sequence to pooled estimates from multiple CS scans collected in succession from a mixed sample of healthy young and older participants as well as symptomatic patients with MCI or dementia likely due to AD or FTLD. We found that pooling estimates from four CS scans (total acquisition time = 4’48’’) yields an average 34% reduction in measurement error compared with estimates based on a single ADNI scan (Elliott et al., 2024). Importantly, the magnitude of error reductions by pooling estimates was comparable across healthy young adults (33%), healthy older adults (34%), and AD/FTLD patients (35%). Taken together, these findings show that cluster scanning improves morphometric precision by reducing measurement error and its utility is clearly demonstrated in patient populations where poorer data quality may be expected. It remains unclear, however, whether the increased measurement precision offered by cluster scanning will translate to enhanced sensitivity to detect subtle longitudinal morphometric change over short intervals within individual patients.

In this pilot study, we performed a proof-of-concept investigation of the utility of cluster scanning to detect short-interval longitudinal cortical atrophy within individual patients diagnosed with MCI or mild dementia likely due to AD or FTLD. At the time of study execution, some of the accelerated MRI sequences above were not yet readily available. Therefore, each patient (*N* = 4) underwent repeated fast low-resolution (1.2 mm) multi-echo MPRAGE (MEMPRAGE) acquisitions (Holmes et al., 2015; Mair et al., 2012) across three time points over a 6-month period. We then calculated the rate of longitudinal atrophy in each patient over 3-month and 6-month intervals based on pooled morphometric estimates obtained from up to 32 scans per time point. We hypothesized that longitudinal cortical atrophy would be detectable over 3- and 6-month intervals within individual patients with AD or FTLD, thus demonstrating the *sensitivity* of cluster scanning to longitudinal short-interval atrophy. Furthermore, we hypothesized that the magnitude of longitudinal cortical atrophy would be greater in phenotypically more vulnerable (i.e., core atrophy) regions than in less vulnerable regions uniquely defined in each patient with AD or FTLD, thus demonstrating the *specificity* of cluster scanning to longitudinal short-interval atrophy in core regions.

## 2. Methods

### 2.1. Participants

Four participants with MCI or mild dementia (Clinical Dementia Rating, CDR = 0.5 or 1) likely due to AD (*n* = 3; with positive amyloid, tau, and neurodegeneration imaging biomarkers) (Jack et al., 2016) or FTLD (*n* = 1) were recruited through the Massachusetts General Hospital Frontotemporal Disorders Unit. Of the three AD patients, two of them clinically presented with logopenic variant Primary Progressive Aphasia (lvPPA) (Gorno-Tempini et al., 2011), while one presented with the Posterior Cortical Atrophy (PCA) syndrome (Crutch et al., 2017; Mendez et al., 2002; Tang-Wai et al., 2004). The FTLD patient’s presentation was semantic variant PPA (svPPA) (Gorno-Tempini et al., 2011). We chose this group of participants to explore the viability of longitudinal cluster scanning across individuals with distinct patterns and etiologies of atrophy (Gorno-Tempini et al., 2004; Mummery et al., 2000; Whitwell et al., 2007). CDR global, CDR Sum-of-Boxes (CDR-SB), and FTLD-adapted CDR-SB (FTLD-CDR-SB) scores (Knopman et al., 2008) were obtained from recent clinical or research visits. See **Table 1** for demographic and clinical characteristics of the sample. The participants self-reported that their ethnicity and race were non-Hispanic Caucasian.

**Table 1.** Demographic and clinical characteristics of the sample.

| Participant ID | lvPPA-1 | lvPPA-2 | PCA | svPPA |
| --- | --- | --- | --- | --- |
| Likely etiology | AD | AD | AD* | FTLD** |
| Sex | M | M | F | M |
| Age group at baseline | 65-69 | 70-74 | 55-59 | 70-74 |
| Years of education | 20 | 20 | 16 | 18 |
| CDR global | 1 | 0.5 | 0.5 | 0.5 |
| CDR-SB | 4 | 3 | 3 | 0.5 |
| FTLD-CDR-SB | 5 | 4 | 3 | 1 |

Notes: AD = Alzheimer's Disease; FTLD = Frontotemporal Lobar Degeneration; CDR = Clinical Dementia Rating; CDR-SB = Clinical Dementia Rating Sum-of-Boxes; lvPPA = logopenic variant Primary Progressive Aphasia; svPPA = semantic variant Primary Progressive Aphasia; PCA = Posterior Cortical Atrophy. \*autopsy-proven AD pathology; \*\*autopsy-proven FTLD TDP43 pathology.

### 2.2. MRI data acquisition

MRI data used to assess longitudinal atrophy in each participant were collected at the Harvard Center for Brain Science using a 3T Siemens MAGNETOM Prisma^fit^ MRI scanner (Siemens Healthineers AG; Erlangen, Germany) and the vendor’s 64-channel head coil. Specifically, we utilized a fast T1w MEMPRAGE sequence: Acquisition time = 2’23’’, repetition time [TR] = 2.2 s, echo times [TEs] = 1.57/3.39/5.21/7.03 ms, flip angle = 7°, inversion time [TI] = 1.1 s, slice orientation = sagittal, matrix size = 192 × 192 × 144, in-plane generalized auto-calibrating partial parallel acquisition (GRAPPA) acceleration factor = 4, voxel size = 1.2 mm isotropic voxels (Holmes et al., 2015; Mair et al., 2012). This sequence has been shown to yield morphometric estimates that are highly consistent (*R*^2^ > 0.9 for several gray and white matter structures) with those obtained from more conventional MPRAGE scans with longer acquisition time and higher spatial resolution (Mair et al., 2012). All fast structural MRI scans were acquired with motion-tracking enabled via volumetric navigators (vNavs) (Reuter et al., 2015; Tisdall et al., 2016) to provide estimates of head motion in each scan, although prospective motion correction was not performed. During the scanning sessions, participants were encouraged to remain still and given the option to watch video clips (e.g., a nature documentary) or to listen to music. Inflatable cushions were used to provide additional hearing protection and to immobilize the participants’ heads. Every 5-10 minutes participants were given feedback about motion and reminded to stay still.

The cluster scanning protocol comprised up to 32 fast structural MRI scans at each of the three timepoints (baseline, 3-month follow-up, and 6-month follow-up). At each timepoint, participants completed four scanning sessions over the course of two separate days (mean interval = 4.5 ± 1.88 days), with two sessions each day. In the first session of the day, eight fast MEMPRAGE scans were collected along with additional structural and functional acquisitions (not analyzed here). Once complete, each participant was taken out of the scanner for a brief break to stretch and use the restroom if needed. Following the break, the second session of the day was conducted where each participant was repositioned, the scanner re-shimmed, and the same protocol repeated, including another set of eight fast MEMPRAGE scans. Altogether, each visit consisted of two scanning sessions with a break in between lasting no more than 2 hours. Due to technical difficulties, we lost half of the scans acquired from the PCA participant at baseline.

We additionally acquired from each participant a standard MEMPRAGE scan at the MGH Athinoula A. Martinos Center for Biomedical Imaging using a 3T Siemens Tim Trio MRI scanner and the vendor’s 12-channel head coil: Acquisition time = 5’53’’, TR = 2.53 s, TEs = 1.64/3.5/5.36/7.22 ms, flip angle = 7°, TI = 1.2 s, slice orientation = sagittal, matrix size = 256 × 256 × 176, GRAPPA acceleration factor = 2, voxel size = 1 mm isotropic voxels. These data were collected as part of their participation in different studies within 1 year preceding the baseline of the present study (mean time before baseline = 7.7 ± 4.7 months) and were used to independently define regions of interest (ROI) masks for the analysis of longitudinal cortical atrophy over 3- and 6-month periods (see *ROI mask creation* below).

### 2.3. MRI data preprocessing

All structural MRI data were processed with FreeSurfer version 6.0.0 using the recon-all pipeline, which involved intensity normalization, skull stripping, and an automated segmentation of cerebral white matter to locate the gray matter/white matter boundary. Defects in the surface topology were corrected (Fischl et al., 2001), and the gray/white boundary was deformed outward using an algorithm designed to obtain an explicit representation of the pial surface. We visually inspected each participant’s cortical surface reconstruction for technical accuracy. Cortical thickness was calculated as the closest distance from the gray/white boundary to the gray/cerebrospinal fluid boundary at each vertex on the tessellated surface (Fischl & Dale, 2000). We resampled each participant’s native-space cortical thickness maps to *fsaverage* space (with ∼160k vertices per hemisphere) and geodesically smoothed these resampled thickness maps with FWHM = 15 mm.

We examined head motion estimates calculated by vNavs for fast structural MRI scans obtained in clusters. During each TR, the vNavs system estimates the participant’s head displacement relative to the first TR of the scan. To summarize head motion, we computed the root mean square (RMS) of the displacement over all points inside a sphere with 64 mm radius, initially centered at isocenter (Jenkinson, 1999), resulting in one summary metric of estimated head motion per TR. The framewise motion estimates were then averaged over the duration of the scan, which yielded a single motion score per scan, RMS displacement per minute (RMSpm). In this study, we discarded from each participant all scans with RMSpm > 10 mm/min. The number of scans discarded based on this criterion for each participant was: lvPPA-1 = 5 (∼5%), lvPPA-2 = 1 (∼1%), PCA = 13 (∼16%), svPPA = 0 (0%).

### 2.4. ROI mask creation

Given that our participants were in the early clinical stages of AD or FTLD, we hypothesized that they would continue to exhibit longitudinal atrophy in cortical regions where some atrophy had already been present prior to their participation in this study. To identify these phenotypically vulnerable cortical regions tailored to the idiosyncratic anatomy of each participant, we first processed the standard structural MRI scan acquired independently of the cluster data following the procedure described above. We then converted whole-cortex vertex-wise estimates of cortical thickness to *W*-scores (Jack et al., 1997; Katsumi et al., 2023). *W*-scores are analogous to *Z*-scores adjusted for specific covariates of no interest, which in this study were participants’ age and sex. Separately for each vertex, we first performed a multiple linear regression analysis using cortical thickness data obtained from a group of biomarker-confirmed amyloid-negative (Aβ-) control participants (mean age = 67.4 ± 4.8, 13 men/12 women) (Katsumi et al., 2023), with the following equation:

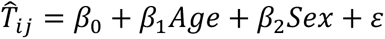

where 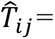 the predicted cortical thickness at vertex i and for Aβ- control participant j. This regression model produced beta coefficient values for age and sex and individual values of residuals. Using these parameters, we then computed *W*-scores for each vertex and participant with the following formula:

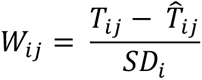

where *T*_*ij*_= the observed cortical thickness at vertex i and for participant j, 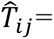 the predicted cortical thickness at vertex i and for patient j based on age and sex of the participant and beta coefficients obtained from Aβ- controls, and *SD*_*i*_ = the standard deviation of the individual residuals obtained from Aβ- controls for vertex i. Because *W*-scores in this study were calculated using cortical thickness, more negative values indicate greater cortical atrophy relative to what would be expected solely based on age and sex of each participant.

For each participant, we deployed a variant of the individualized cortical thinning signature (iCORTS) approach that we previously developed (Bakkour et al., 2008). Specifically, we defined phenotypically vulnerable (“core” atrophy) ROIs by first thresholding their *W*-score atrophy map at *W* < −2. Using FreeSurfer’s tksurfer tools, we visually inspected the thresholded *W*-score map then manually identified the clusters that anatomically correspond to cortical regions where prominent neurodegeneration is typically observed in each AD or FTLD clinical syndrome (Crutch et al., 2017; Gorno-Tempini et al., 2011; Gorno-Tempini et al., 2004). In two of the four participants, we additionally used anatomical parcellation labels generated by FreeSurfer (Desikan et al., 2006) to facilitate anatomical localization of core ROIs. Furthermore, we identified cortical regions where minimal atrophy was present and hence comparatively less longitudinal atrophy would be expected (“control” ROIs). We initially defined these ROIs as the precentral and postcentral gyri (i.e., primary motor and somatosensory cortex, respectively) based on each participant’s individualized parcellation of these areas generated by FreeSurfer, as they are typically relatively spared in patients with lvPPA, svPPA, and PCA. We then thresholded each participant’s *W*-score atrophy map at *W* < −0.25 and subtracted any suprathreshold vertices from the precentral/postcentral gyrus labels to further minimize the presence of atrophy within these regions. The final ROI masks used for statistical analysis of each participant’s fast structural MRI data are shown in **Figure 2**. For each fast structural MRI scan, mean cortical thickness of each ROI was calculated by averaging thickness values at all vertices falling within its boundaries.

**Figure 2.**
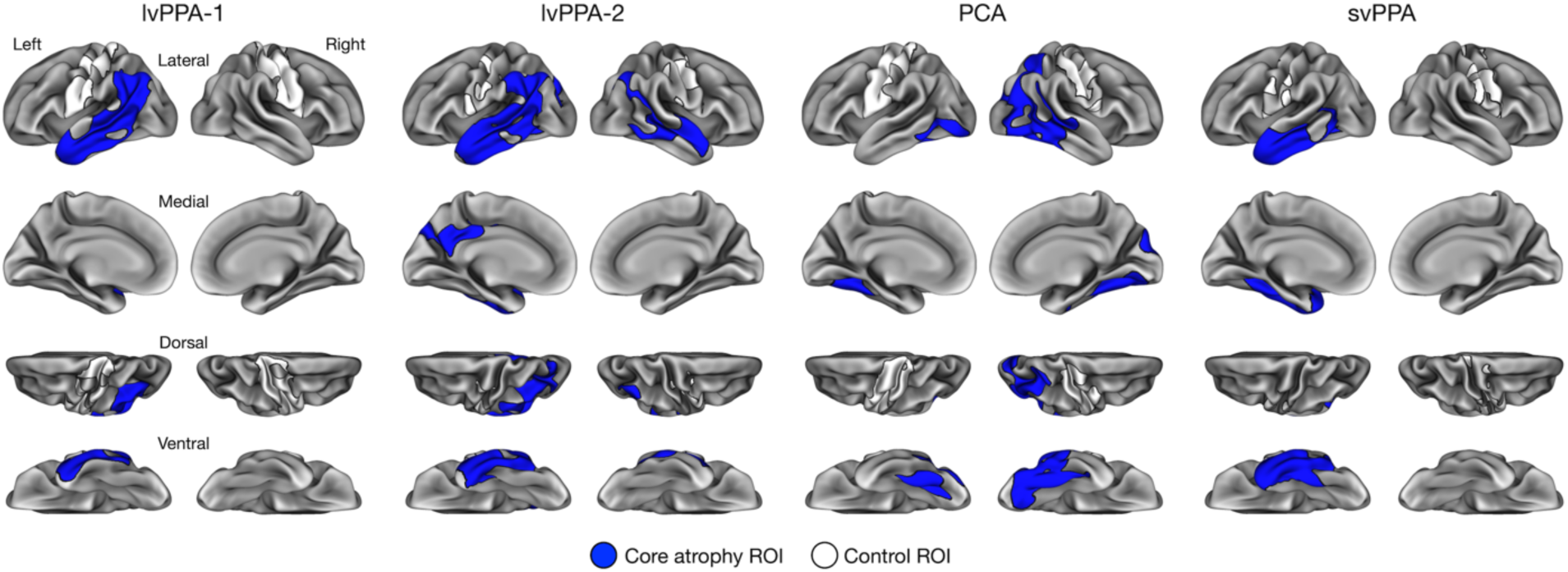
Individualized cortical thinning signatures (iCORTS) ROI masks. For each participant, core atrophy ROI (blue) and control ROI (white) masks were defined based on a standard structural MRI scan collected prior to their participation in the current study (see *2.4. ROI mask creation*).

### 2.5. Statistical analysis

To examine longitudinal cortical atrophy, we constructed linear mixed-effects models separately for each individual participant using the *lme* function from the *nlme* package (version 3.1-164) (Pinheiro et al., 2023) run on *R* version 4.2.1. These models were fit by maximizing the restricted likelihood and included Timepoint (baseline, 3 months, and 6 months) and ROI (core and control) as fixed predictors of interest, estimated head motion (RMSpm) as a covariate of no interest, and nested random intercepts for scanning session within day. We additionally included variance weights to allow for heteroscedasticity in cortical thickness estimates between core atrophy and control ROIs across timepoints.

To test our first hypothesis regarding the sensitivity of cluster scanning, we examined the main effect of Timepoint estimated by each model. Planned comparisons investigated pairwise differences in cortical thickness separately for core atrophy and control ROIs using the *emmeans* package (version 1.10.0). To test our second hypothesis regarding the specificity of cluster scanning, we examined the interaction effect between Timepoint and ROI. If a significant interaction was identified, we computed the magnitude of longitudinal atrophy between a given pair of timepoints separately for core atrophy and control ROIs and compared these estimates. *p*- values were Bonferroni-corrected for multiple statistical comparisons within each participant based on the number of contrasts in question.

Additionally, we performed an exploratory vertex-wise analysis of longitudinal cortical atrophy within the boundaries of each participant’s core atrophy ROIs. This analysis allowed us to examine potential subregional specificity in the localization of longitudinal atrophy and how it might change over different time intervals. Using vertex-wise cortical thickness data as inputs, we constructed linear mixed-effects models separately for each vertex within each participant’s core atrophy ROIs. Given the number of vertices involved in this analysis, it is possible that the maximal random-effects structure might be too complex for some of them, resulting in a singular fit. To overcome this potential issue, we initialized model estimation for all vertices with the nested random intercepts for scanning session within day as described above. In the case of a singular fit, the model was refit after dropping a random intercept for scanning session. If this model also resulted in a singular fit, a fixed effects only model was fit to the data. Using vertex-wise estimated marginal means, we calculated surface maps representing the percent change in cortical thickness from one timepoint to another for the three pairwise contrasts: Baseline vs. 3 months, 3 months vs. 6 months, and Baseline vs. 6 months. The magnitude of change was considered statistically significant at each vertex if *p* < .05 for the main effect of Timepoint and *p* < .017 for the corresponding pairwise contrast (Bonferroni-corrected for the number of comparisons). Due to the exploratory nature of this analysis, we did not otherwise adjust *p*-values for the number of vertices within each participant’s ROIs.

## 3. Results

### 3.1. Cluster scanning detects longitudinal cortical atrophy over 3- and 6-month intervals in individual patients with AD or FTLD

In all four participants, we found a significant main effect of Timepoint (all *p*’s < .0001). Planned comparisons revealed that, in all participants, core regions showed longitudinal atrophy from baseline to 3 months (lvPPA-1: *b* = 0.032, *SE* = 0.004, *t*(172) = 3.47, *p* < .0001; lvPPA-2: *b* = 0.026, *SE* = 0.004, *t*(178) = 6.25, *p* < .0001; PCA: *b* = 0.034, *SE* = 0.004, *t*(124) = 7.68, *p* < .0001; svPPA: *b* = 0.035, *SE* = 0.006, *t*(182) = 6.03, *p* < .0001) and from baseline to 6 months (lvPPA-1: *b* = 0.056, *SE* = 0.004, *t*(172) = 12.90, *p* < .0001; lvPPA-2: *b* = 0.034, *SE* = 0.004, *t*(178) = 8.82, *p* < .0001; PCA: *b* = 0.041, *SE* = 0.004, *t*(124) = 9.25, *p* < .0001; svPPA: *b* = 0.044, *SE* = 0.006, *t*(182) = 7.58, *p* < .0001) (**Figure 3**). Only the lvPPA-1 participant showed longitudinal atrophy in core regions from 3 months to 6 months follow-up at a corrected statistical threshold: *b* = 0.023, *SE* = 0.004, *t*(172) = 5.38, *p* < .0001. In three participants, control regions showed longitudinal atrophy from baseline to 6 months lvPPA-1: *b* = 0.019, *SE* = 0.006, *t*(172) = 3.03, *p* < .003; lvPPA- 2: *b* = 0.034, *SE* = 0.013, *t*(178) = 2.74, *p* < .007; PCA: *b* = 0.033, *SE* = 0.009, *t*(124) = 5.14, *p* < .0001). Only the PCA participant showed significant atrophy in control regions from 3 months to 6 months follow-up at a corrected statistical threshold: *b* = 0.039, *SE* = 0.008, *t*(124) = 5.14, *p* < .0001. **Table 2** summarizes mean cortical thickness for each ROI, timepoint, and participant estimated by our linear mixed-effects models, whereas **Table 3** summarizes fixed effects parameters estimated by these models.

**Figure 3.**
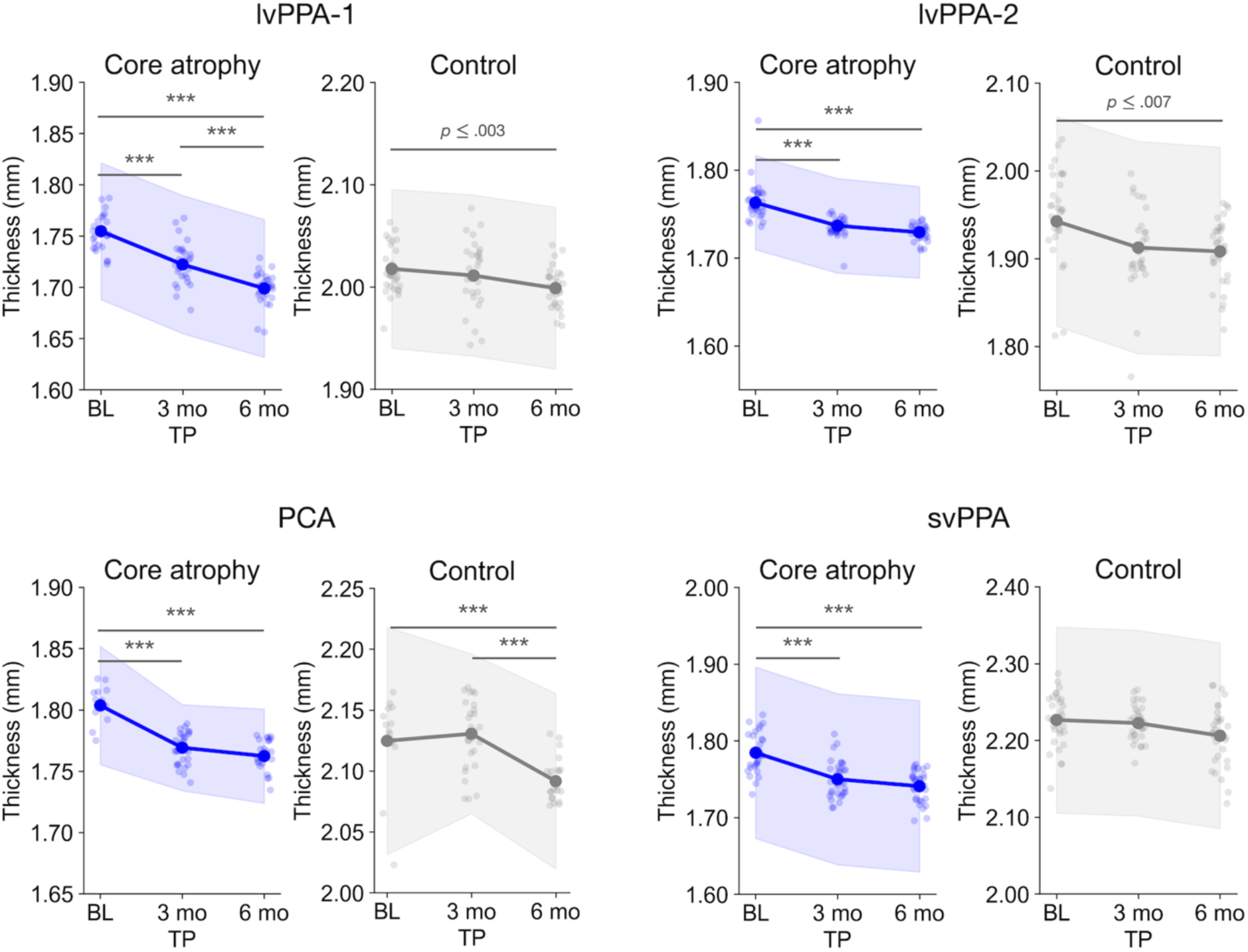
Pooling morphometric estimates via cluster scanning reveals short-interval longitudinal cortical atrophy in neurodegenerative patients. Each plot displays estimated marginal means for cortical thickness at each time point (larger opaque points) obtained from linear mixed-effects models, overlaid on top of raw thickness values derived from individual fast structural MRI scans (smaller transparent points). Shaded bands represent 95% confidence intervals. Statistical significance is denoted only if a given comparison survives a Bonferroni-corrected threshold of *p* < .008 (*p*_bonf_ = .05/6). \*\*\**p* < .0001. TP = Timepoint; BL = baseline; 3 mo = 3 months; 6 mo = 6 months.

**Table 2.** Estimated marginal means for cortical thickness by participant, region of interest, and timepoint.

| Participant ID | ROI | Timepoint |  |  |
| --- | --- | --- | --- | --- |
|  |  | Baseline | 3 months | 6 months |
| lvPPA-1 | Core | 1.75 (0.0053) | 1.72 (0.0053) | 1.70 (0.0053) |
|  | Control | 2.02 (0.0061) | 2.01 (0.0062) | 2.00 (0.0062) |
| lvPPA-2 | Core | 1.76 (0.0042) | 1.74 (0.0042) | 1.73 (0.0041) |
|  | Control | 1.94 (0.0095) | 1.91 (0.0095) | 1.91 (0.0093) |
| PCA | Core | 1.80 (0.0038) | 1.77 (0.0028) | 1.76 (0.0030) |
|  | Control | 2.12 (0.0073) | 2.13 (0.0052) | 2.09 (0.0057) |
| svPPA | Core | 1.78 (0.0088) | 1.75 (0.0088) | 1.74 (0.0088) |
|  | Control | 2.23 (0.0095) | 2.22 (0.0095) | 2.21 (0.0095) |

Notes: Values indicate estimated marginal means (standard error of the mean).

**Table 3.**
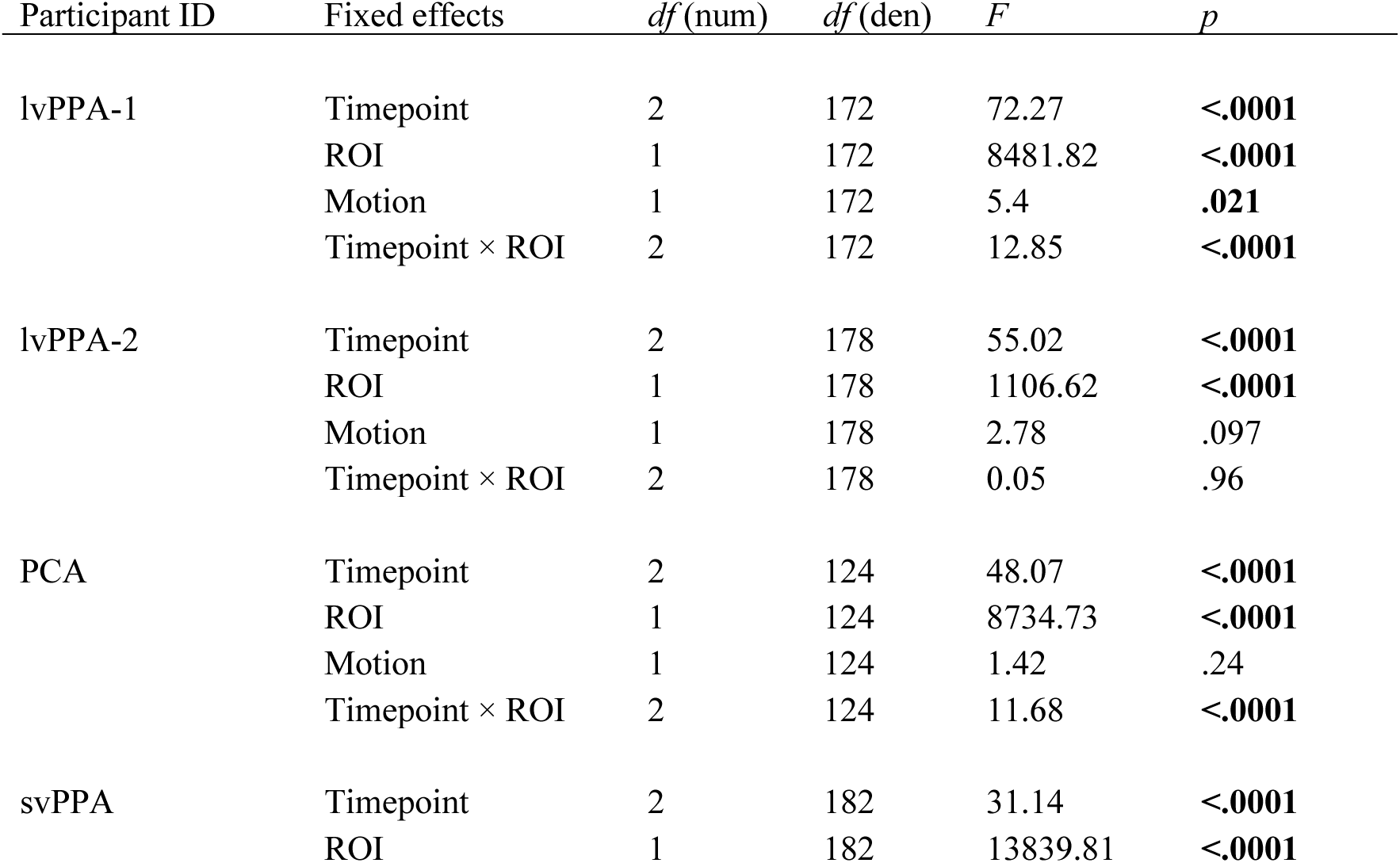
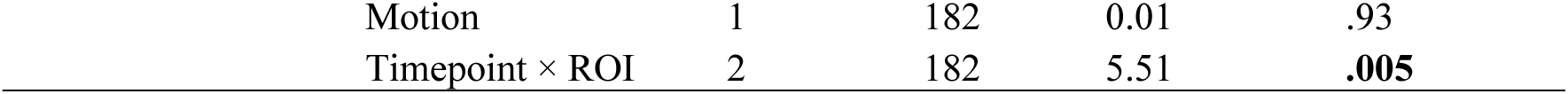
Estimated fixed effects parameters from linear mixed-effects models.

Next, we tested the critical interaction between the core and control ROIs to ask whether atrophy was preferential to the hypothesized vulnerable regions. In three out of four participants (lvPPA-1, PCA, svPPA), we found a significant interaction between Timepoint and ROI (**Table 3**). In each of the three participants, we compared the magnitude of longitudinal atrophy (i.e., a decrease in thickness) from a given timepoint to another between core and control regions. For two participants (lvPPA-1 and svPPA), this interaction effect was driven by core regions exhibiting longitudinal atrophy of a greater magnitude from baseline to 3 months (lvPPA-1: *b* = Δ0.026, *SE* = 0.007, *t*(172) = 3.47, *p* ≤ .0001; svPPA: *b* = Δ0.03, *SE* = 0.01, *t*(182) = 3.18, *p* ≤ .002) and from baseline to 6 months (lvPPA-1: *b* = Δ0.037, *SE* = 0.008, *t*(172) = 4.91, *p* < .0001; svPPA: *b* = Δ0.023, *SE* = 0.01, *t*(182) = 2.42, *p* ≤ .017) compared with control regions. For the PCA participant, core regions showed greater longitudinal atrophy than control regions from baseline to 3 months as predicted (*b* = Δ0.04, *SE* = 0.01, *t*(124) = 4.14, *p* < .0001). However, this difference was not significant from baseline to 6 months (*b* = Δ0.008, *SE* = 0.01, *t*(124) = 0.82, *p* ≤ .41), which was driven by control regions unexpectedly exhibiting longitudinal atrophy of a larger magnitude from 3 months to 6 months follow-up compared with core regions (*b* = Δ-0.032, *SE* = 0.008, *t*(124) = - 3.89, *p* < .0001).

### 3.2. Vertex-wise analysis reveals the possible spatiotemporal trajectory of short-interval longitudinal cortical atrophy

We performed an exploratory vertex-wise analysis of longitudinal cortical atrophy within the boundaries of core atrophy regions in each participant. This analysis identified prominent longitudinal atrophy within each participant’s phenotypically vulnerable cortical regions from baseline to 3 months and from baseline to 6 months, with modest atrophy observed between 3 months and 6 months, consistent with ROI-based results reported above. In lvPPA-1, we found longitudinal atrophy from baseline to 3 months prominently in the anterior lateral temporal lobe and inferior parietal lobule, with additional involvement of the caudal middle temporal gyrus and posterior inferior parietal lobule from 3 months to 6 months. By 6 months, longitudinal atrophy was detectable throughout core regions (**Figure 4**). In lvPPA-2, we identified left-lateralized longitudinal atrophy primarily in the anterior temporal lobe and anterior inferior parietal lobule similar to lvPPA-1, with minimal changes observed from 3 months to 6 months. By 6 months, longitudinal atrophy became more spatially extensive in the lateral portion of the ROI, although the medial parietal cortex—despite being part of the ROI mask—showed minimal atrophy over 6 months (**Figure 5**). In PCA, we found longitudinal atrophy from baseline to 3 months localized to ventral and lateral temporo-occipital areas, with minimal changes from 3 months to 6 months. By 6 months, longitudinal atrophy was slightly more pronounced in the right middle and inferior temporal gyri, but the overall spatial extent of atrophy was rather focal relative to the total area of the ROI mask (**Figure 6**). Finally, in svPPA, we identified longitudinal atrophy from baseline to 3 months localized to the left temporal pole/anterior temporal lobe, with minimal changes in the middle temporal gyrus from 3 months to 6 months. By 6 months, longitudinal atrophy was greater in its extent, covering most areas of the left-lateralized ROI mask (**Figure 7**).

**Figure 4.**
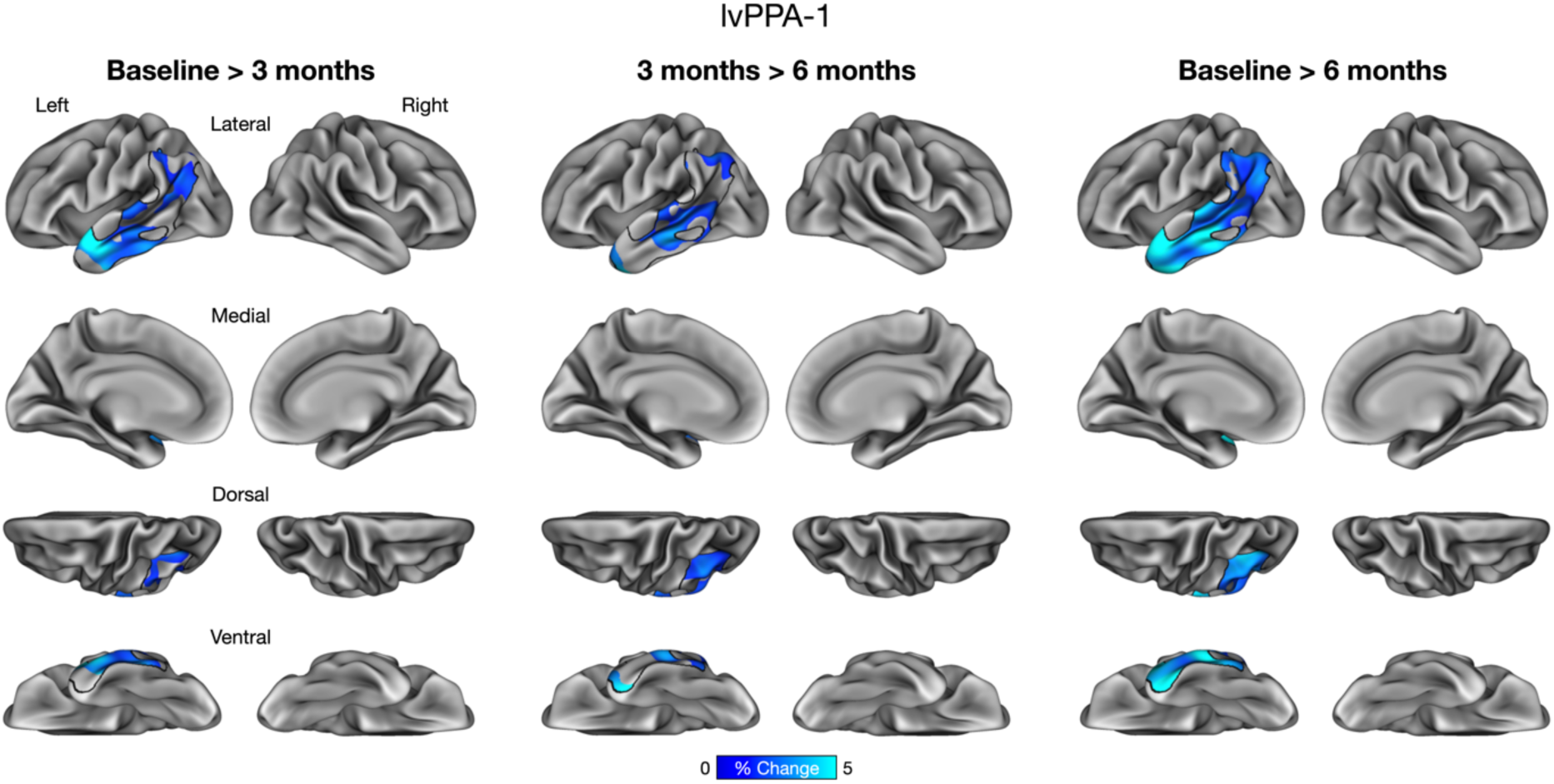
Vertex-wise analysis of longitudinal cortical atrophy within core atrophy ROIs in a patient with logopenic variant Primary Progressive Aphasia (lvPPA-1).

**Figure 5.**
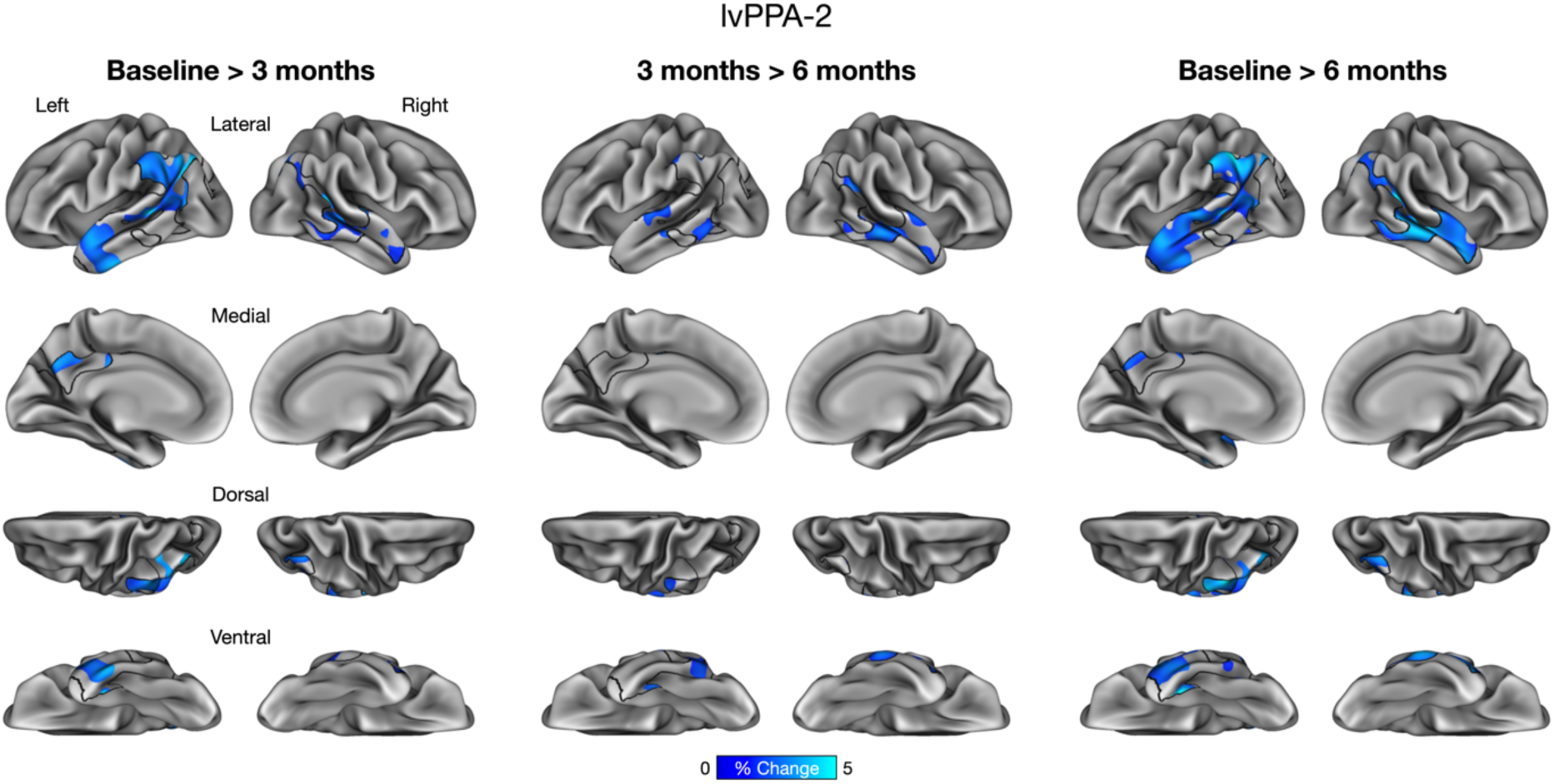
Vertex-wise analysis of longitudinal cortical atrophy within core atrophy ROIs in a patient with logopenic variant Primary Progressive Aphasia (lvPPA-2).

**Figure 6.**
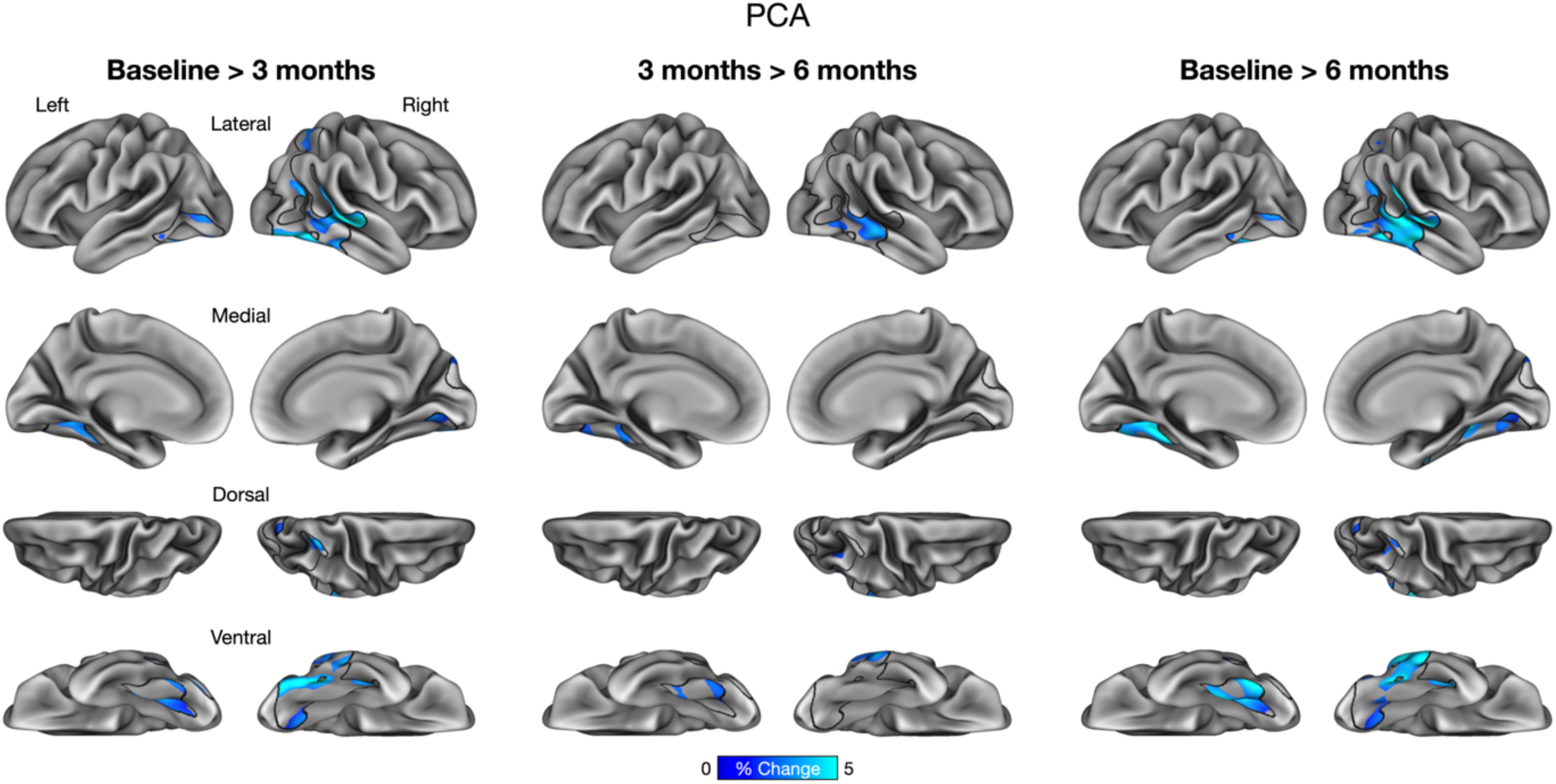
Vertex-wise analysis of longitudinal cortical atrophy within core atrophy ROIs in a patient with Posterior Cortical Atrophy (PCA).

**Figure 7.**
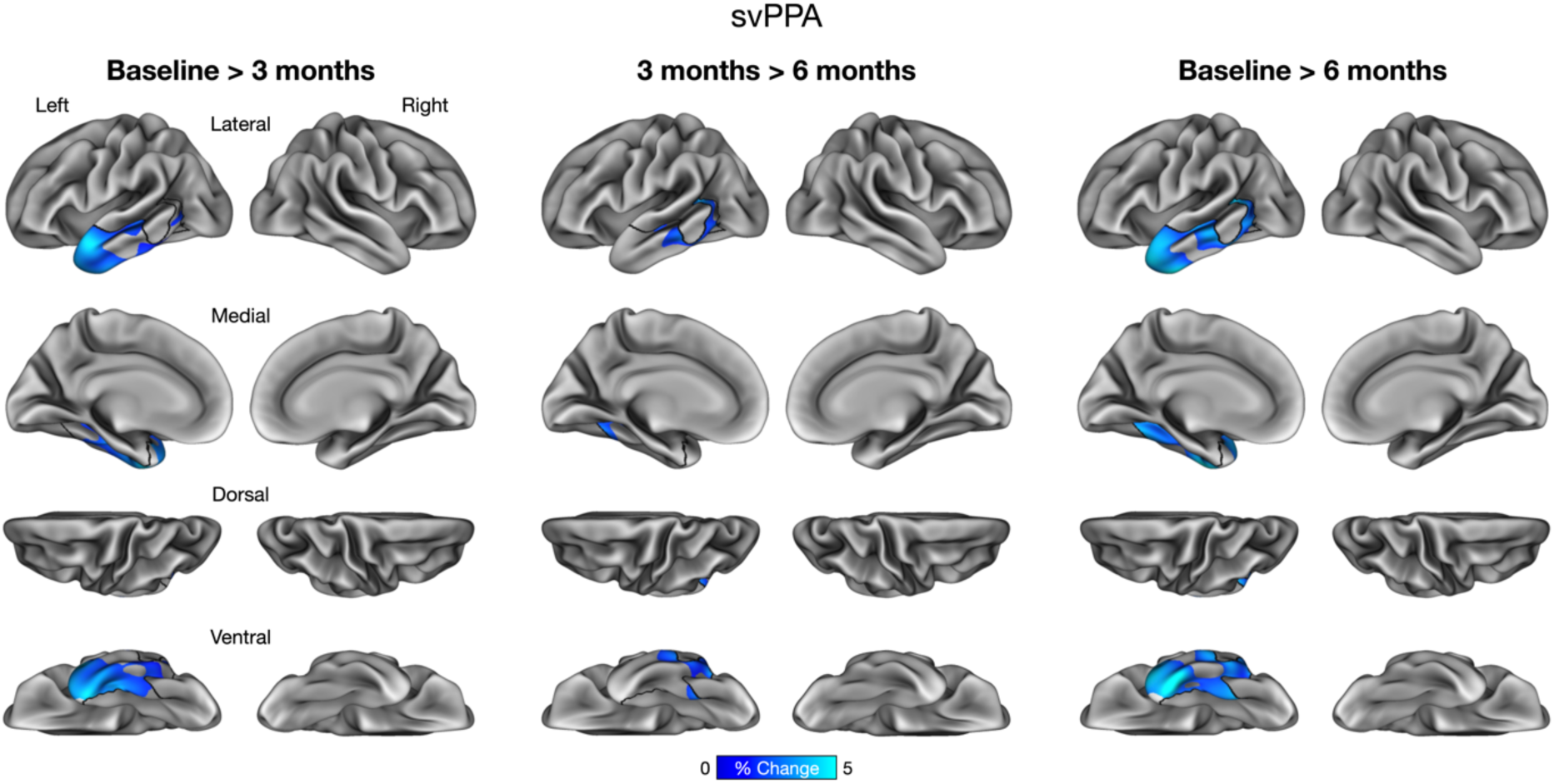
Vertex-wise analysis of longitudinal cortical atrophy within core atrophy ROIs in a patient with semantic variant Primary Progressive Aphasia (svPPA).

## 4. Discussion

The goal of this study was to evaluate the utility of dense sampling of structural MRI data at each timepoint—cluster scanning—in the measurement of longitudinal cortical atrophy over short time intervals by means of increasing brain morphometric precision. In each individual patient with AD or FTLD, we found that phenotypically vulnerable cortical (“core atrophy”) regions exhibited longitudinal atrophy that was detectable over 3- and 6-month intervals. Thus, this novel structural MRI measure is sensitive to the detection of progressive neurodegeneration in intervals that are half or even less than half of those typically used in large, multicenter natural history and biomarker studies such as ADNI, DIAN, ALLFTD, LEADS, PPMI, and many others. In three of four participants, the magnitude of longitudinal atrophy in core regions was statistically greater than the magnitude of atrophy observed in control regions over 6 months (*n* = 2) and at least one of the 3-month intervals (*n* = 3), which additionally demonstrates the specificity of cluster scanning in detecting preferential longitudinal atrophy in vulnerable regions over these intervals. An exploratory vertex-wise analysis revealed subregional specificity in the localization of longitudinal atrophy within phenotypically vulnerable regions, the extent of which was variable across individual participants. These findings provide proof-of-concept evidence demonstrating the utility of MRI cluster scanning for the detection of progressive atrophy over intervals shorter than those typically used in longitudinal studies of neurodegenerative dementias.

The current results support our initial hypothesis and suggest that cortical regions that had been already exhibiting atrophy in the early symptomatic stages of these patients’ neurodegenerative cognitive impairment continued to do so over time. This is consistent with prior work identifying partial spatial overlap in the localization of baseline atrophy and longitudinal atrophy among individuals with lvPPA, PCA, and svPPA who were mildly impaired at baseline (Kumfor et al., 2016; Rohrer et al., 2013; Sintini et al., 2020). Surprisingly, although we detected atrophy over the first 3-month interval, we did not find strong evidence of continued progressive atrophy in our participants’ phenotypically vulnerable regions as a whole from 3 months to 6 months. It is possible that some areas within these regions had become saturated with neurodegenerative changes, thus reducing the overall magnitude of cortical atrophy detectable during this period. Our exploratory vertex-wise analysis provided evidence partially in support of this possibility, revealing an idiosyncratic spatiotemporal trajectory of cortical atrophy progression within each participant. In addition, our findings suggest that the time-dependent changes in cortical thickness do not represent a global phenomenon, as indicated by longitudinal atrophy of greater magnitudes in core regions than in control regions observed in the majority of the participants.

The vertex-wise spatial topography of longitudinal atrophy in the patient with PCA was focal relative to the total area of her core atrophy ROI. One possibility is that, by the time baseline MRI assessments were conducted for this study, atrophy had already spread to other areas of the cerebral cortex in this participant. This seems unlikely, however, given that this participant was at the MCI stage throughout her participation in this study and that PCA patients with similar levels of impairment have been shown to exhibit longitudinal atrophy in temporo-occipital regions consistent with this participant’s core atrophy ROI (Sintini et al., 2020). It is also possible that this was a technical issue, since half the data acquired from this participant at baseline were unavailable for analyses, and she also exhibited the highest rate of data exclusion (16%) due to excessive head motion. As such, results based on the PCA participant’s data should be interpreted with caution.

Beyond the proof-of-concept evidence that supports further investigation of the cluster scanning approach, several questions remain. First, it will be important for future studies to determine how many scans per timepoint are required to reliably estimate longitudinal atrophy over short intervals. Although low resolution structural MRI scans employed in this study can be acquired more quickly than conventional 1 mm-isotropic MRI scans, collecting 32 of them per timepoint over two days would significantly increase participant burden and study cost. Such a design would also be impractical in typical neuroimaging studies. In this context, cluster scanning using extremely rapid sequences, including CS scans (acquisition time = 1’12’’), will be helpful in boosting morphometric precision necessary for the detection of short-interval atrophy while maintaining a reasonable scan duration. We have recently demonstrated that longitudinal atrophy can be robustly detected in patients with cognitive impairment and in some cognitively unimpaired older adults over one year using clusters of eight CS scans (just under 10 minutes) (Elliott et al., 2025).

Second, future work should investigate the pattern of short-interval longitudinal atrophy in more diverse samples, including individuals with different clinical phenotypes of AD/FTLD (e.g., early-onset AD, behavioral variant FTD) and disease stages, where the magnitude and spatial extent of cortical atrophy may increase at variable rates (Cho et al., 2013; E. Gordon et al., 2021; Staffaroni et al., 2020). Studying such individuals over a large number of short intervals (e.g., every 3-6 months for 2-4 years) would help better understand the heterogeneity in disease progression over finer temporal scales in the context of differences in the topography of neuroanatomical abnormalities and in the molecular drivers of neurodegeneration. In addition, it will be important to examine age-matched, cognitively unimpaired participants in which the rate of longitudinal atrophy over such intervals is usually much smaller in magnitude compared with that detectable in neurodegenerative patients. Establishing this reference based on control data, as we are beginning to do (Elliott et al., 2025), would facilitate the interpretation of the magnitude of longitudinal atrophy in patients and would also be useful in accounting for age dependency in the observed effects.

Third, future studies should also perform external validation of short-interval longitudinal atrophy as measured by cluster scanning against other biomarkers that are used in natural history studies and clinical trials, including amyloid PET, tau PET, ^18^F-fluorodeoxyglucose PET, and plasma and cerebrospinal fluid measures. Clarifying the relationships among these biomarkers would be essential in evaluating and predicting the trajectory of neurodegenerative change in individual patients. For instance, it would be beneficial to understand how the level of cerebral tau accumulation or neurofilament light predicts the rate of cortical atrophy over the next several months. This information may be useful in estimating the rate of downstream cognitive and functional decline, particularly among individuals following an aggressive clinical course.

Finally, this method could be helpful in refining our understanding of the effects of amyloid-reducing monoclonal antibodies on regional brain volume, recognizing the counterintuitive findings to date regarding reductions in brain volume with amyloid-lowering therapies (Belder et al., 2024). We would hope that a comparison of short-interval cluster scanning-based morphometric measurements in cortical areas of high versus low baseline amyloid burden would clarify whether MRI volumetric reductions are specifically localized to areas where amyloid is being removed and not other areas, supporting the concept of pseudo-atrophy.

In sum, the present findings strongly suggest the feasibility of robustly detecting longitudinal cortical atrophy over short intervals within individual patients with suspected AD or FTLD via MRI cluster scanning. These findings have implications for the development of small, early phase, or even personalized (“*N*-of-1”) clinical trials examining disease-modifying effects of therapeutic interventions. Specifically, using cluster scanning, it is conceivable to estimate the natural progression of neurodegeneration in an individual patient across three timepoints over a 6- month run-in phase, followed by an intervention phase that involves MRI assessments every three months for the duration of the trial to precisely identify the timing of its effect on slowing the rate of neurodegeneration. If successful, such a design could dramatically alter the landscape of clinical trials, while also potentially supporting the evaluation of crossover to different treatments (e.g., as in platform trials) to offer patients the maximal opportunity for benefit and to gain the most knowledge from each individual.

## Data and Code Availability

MRI data used in this manuscript may be shared (anonymized) at the request of any qualified investigator for purposes of replicating procedures and results. Data analysis code is available at https://github.com/yutakatsumi/long-cluster-scan.

## Author contributions

Yuta Katsumi: Conceptualization, Data curation, Formal analysis, Investigation, Methodology, Software, Visualization, and Writing – Original Draft. Michael Brickhouse: Data curation, Formal analysis, Investigation, Methodology, Software, and Writing – Original Draft. Lindsay C. Hanford: Data curation, Investigation, and Writing – Review & Editing. Jared A. Nielsen: Data curation, Investigation, and Writing – Review & Editing. Maxwell L. Elliott: Investigation, and Writing – Review & Editing. Ross W. Mair: Investigation, Methodology, Resources, and Writing – Review & Editing. Alexandra Touroutoglou: Investigation and Writing – Review & Editing. Mark C. Eldaief: Investigation and Writing – Review & Editing. Randy L. Buckner: Conceptualization, Data curation, Formal analysis, Funding acquisition, Investigation, Resources, Supervision, and Writing – Review & Editing. Bradford C. Dickerson: Conceptualization, Data curation, Formal analysis, Funding acquisition, Investigation, Resources, Supervision, and Writing – Review & Editing.

## Ethics statement

The study protocol was approved by the Institutional Review Board of Mass General Brigham Healthcare. All participants provided written informed consent in accordance with the guidelines of the Institutional Review Board of Mass General Brigham Healthcare and were compensated.

## Acknowledgments

We greatly appreciate the dedication of the participants in this study and their family members, without whose time and effort this work would not have been possible. This research was supported by US National Institutes of Health (R01 AG081249, R01 DC014296, R01 NS131395, K01 AG084820, R21 AG080588, K23 DC016912, K00 AG068432, P50 AG005134, P30 AG062421), Alzheimer’s Association (AARG-24-1309264), and by the Tommy Rickles Chair in Primary Progressive Aphasia Research. This research was carried out in part at the Athinoula A. Martinos Center for Biomedical Imaging at the MGH, using resources provided by the Center for Functional Neuroimaging Technologies, P41 EB015896, a P41 Biotechnology Resource Grant supported by the National Institute of Biomedical Imaging and Bioengineering (NIBIB), National Institutes of Health. This work also involved the use of instrumentation supported by the NIH Shared Instrumentation Grant Program and/or High-End Instrumentation Grant Program; specifically, grant number(s) S10 RR021110, S10 RR023043, S10 RR023401 (awarded to the Martinos Center for Biomedical Imaging) and S10 OD020039 (awarded to Harvard University Center for Brain Science).

